# Comparison of three linezolid management strategies for peripheral neuropathy in multidrug- or rifampicin-resistant tuberculosis treatment: a target trial emulation

**DOI:** 10.64898/2026.03.14.26348377

**Authors:** Matthew Romo, Allison LaHood, Carole D. Mitnick, Michael L. Rich, Letizia Trevisi, Alena Skrahina, Lawrence Oyewusi, Mathieu Bastard, Palwasha Y. Khan, Helena Huerga, Uzma Khan, Edwin Herrera Flores, Hakob Atshemyan, Catherine Hewison, Mahmud Rashitov, Nazgul Samieva, Camilo Gómez-Restrepo, Aga Krisnanda, Tinatin Kotrikadze, Faisal Siraj, Abdul Wali Khan, Norbert Ndjeka, Malik Adenov, Kwonjune J. Seung, Andargachew Kumsa, Molly F. Franke

## Abstract

**Background:** Peripheral neuropathy frequently leads to linezolid dose reductions or interruptions during multidrug- or rifampicin-resistant tuberculosis (MDR/RR-TB) treatment. The implications of these modifications on treatment success are uncertain.

**Methods:** We conducted a target trial emulation using the endTB Observational Study among individuals who developed non-severe peripheral neuropathy while receiving linezolid 600 mg daily within 6 months of initiating an individualized MDR/RR-TB regimen. We examined three linezolid management strategies: immediate change (i.e., dose reduction, temporary interruption, discontinuation) within Weeks 1–7 after peripheral neuropathy onset, deferred change within Weeks 8–26, and no change (i.e., continuing linezolid 600 mg daily) during Weeks 1–26. To emulate the per-protocol analysis of a trial, we used an approach involving cloning and censoring participant data, and applying inverse probability of censoring weights.

**Results:** Among 303 eligible participants from 12 countries, peripheral neuropathy occurred a median of 11 weeks (interquartile range: 4–18) after treatment initiation. Weighted, standardized probabilities of treatment success were 84.7% (95% CI: 69.2%, 92.9%) for immediate change, 78.9% (95% CI: 65.9%, 87.1%) for deferred change, and 85.2% (95% CI: 80.5%, 89.1%) for no change. Compared with no change, treatment success ratios were 0.99 (95% CI: 0.83, 1.11) for immediate change and 0.93 (95% CI: 0.78, 1.01) for deferred change.

**Conclusions:** We did not find evidence of a substantial negative impact of immediate modification to linezolid on MDR/RR-TB treatment success. Our results support the clinical practice of cautiously adjusting linezolid when needed to manage non-severe peripheral neuropathy.

**Key points:** In this target trial emulation, we found that among individuals with mild or moderate peripheral neuropathy in the first six months of MDR/RR-TB treatment, immediate modifications to linezolid (i.e., dose reduction or interruption) did not substantially compromise treatment success.

## INTRODUCTION

Multidrug- or rifampicin-resistant tuberculosis (MDR/RR-TB) remains a major global health threat, having caused an estimated 150,000 deaths in 2024 [1]. Treatment of this recalcitrant form of TB requires long courses of multiple antibiotics, including linezolid, a repurposed first-in-class oxazolidinone [2-4]. Linezolid is present in all regimens recommended by the World Health Organization (WHO) treatment guidelines, including shorter-duration all-oral regimens [5, 6]. Linezolid’s bactericidal and sterilizing activity results from its inhibition of bacterial protein synthesis. However, off-target binding to human mitochondrial ribosomes can lead to adverse events, such as myelosuppression and neuropathies [7, 8].

Peripheral neuropathy encompasses a range of symptoms, from mild sensory disturbances to disabling pain and motor impairment [9]. It has emerged as one of the main toxicities of MDR/RR-TB treatment [10], and some studies suggest that symptoms can persist long after treatment completion [11, 12]. Linezolid dose is a major determinant of peripheral neuropathy risk [13-16]. Current WHO guidelines recommend a dose of 600 mg daily for most adults but acknowledge that toxicities may necessitate a dose reduction to 300 mg daily [6]. In the first six months of treatment, patients typically present with mild peripheral neuropathy symptoms after at least several weeks on linezolid [10, 17, 18]. Given this timing, peripheral neuropathy remains frequent despite the introduction of 6–9-month regimens [16, 19-23]. New-onset peripheral neuropathy symptoms frequently lead to linezolid modifications, such as dose reduction, temporary interruption, or discontinuation [17, 18, 24, 25]. The course of action must balance managing this adverse event against its potential implications for TB treatment and may change over time as peripheral neuropathy symptoms and TB disease evolve.

Observational cohorts offer an opportunity to evaluate the impact of the timing of linezolid modification on TB treatment outcomes, but are vulnerable to design biases, such as immortal time bias. Immortal time bias arises when patients must, by design, survive long enough to receive a given strategy, making strategies that require longer survival appear advantageous [26]. This is particularly problematic for conditions with high early mortality and loss to follow-up, such as MDR/RR-TB [27]. Target trial emulation, in which observational data are used to mimic a hypothetical randomized trial, can mitigate immortal time and other design biases [28]. Although this approach has been widely used in other fields, it has rarely been applied to TB [29, 30]. Using target trial emulation, we compared the effect of immediate and deferred linezolid modification with continuation of linezolid 600 mg daily on MDR/RR-TB treatment success.

## METHODS

### Data source

We used data from the endTB Observational Study (ClinicalTrials.gov Identifier: NCT03259269), a prospective cohort of people with MDR/RR-TB who received longer (18–24-month) individualized regimens containing bedaquiline and/or delamanid [31]. Study sites in 17 countries enrolled participants from April 2015 through November 2018. The cohort captured routine clinical data under programmatic conditions, with standardized data collection managed centrally. Data collection across sites was prospective and systematic, followed a predefined monitoring schedule, and used standardized forms. Study clinicians made treatment decisions, including regimen selection, monitoring, and management of adverse events, guided by national TB program policies and the endTB clinical guide [32]. Ethics committees of each consortium partner and country approved the study protocol. Participants (or a guardian, if the individual was a minor under local law) provided written informed consent, and older minors provided assent. If individuals were treated multiple times during the study, we included only the first treatment course in these analyses.

At the enrollment visit, participants underwent a full medical history, a clinical examination, laboratory blood testing, chest radiography, sputum smear microscopy and culture, and drug-susceptibility testing [31]. Subsequently, study visits typically occurred at two weeks and then monthly throughout treatment. At follow-up visits, participants had a clinical examination, laboratory blood testing, and sputum smear microscopy and culture. Study staff recorded TB medication orders in a detailed medication log and updated it at each visit. Clinicians assessed peripheral neuropathy using the ACTG Brief Peripheral Neuropathy Screen [33], which is standard in MDR/RR-TB research [19, 20]. Across all sites, this tool was administered routinely as part of the clinical examination at monthly study visits [31]. A diagnosis required a symptom score >0, supplemented by clinical assessment of vibratory sense or deep tendon ankle reflex. Clinicians used the symptom score to assign the adverse event grade. Clinicians followed up adverse events at least monthly to assess changes in severity and need for intervention.

### Target trial specification

The hypothetical target trial would enroll people with pulmonary MDR/RR-TB who initiated a regimen containing linezolid 600 mg daily and had an adverse event of peripheral neuropathy within the first 6 months after treatment initiation (**Table 1**). We focused on this 6-month period because we expected linezolid modifications during the early phase of treatment to have the greatest impact on treatment success [34]. We would exclude participants with severe (grade ≥3) peripheral neuropathy because clinicians would need to take immediate action to manage these patients.

**Table 1.** Components of the target trial specification and how they were emulated using data from the endTB Observational Study.

| Component | Target trial specification | Target trial emulation |
| --- | --- | --- |
| **Eligibility criteria** | <u>Inclusion:</u> Individuals ($\geq 15$ years of age) at eligible sites* with pulmonary MDR/RR-TB who initiated a treatment regimen containing linezolid 600 mg daily<sup>†</sup> and had an adverse event of peripheral neuropathy in the first 6 months after treatment initiation.<br><u>Exclusion:</u> Initial presentation with grade $\geq 3$ peripheral neuropathy. | Same. We also exclude participants who enrolled in the endTB Observational Study $\geq 30$ days since starting MDR/RR-TB treatment to align cohort enrollment with the start of MDR/RR-TB treatment. |
| **Treatment strategies** | Linezolid management strategy in the first 26 weeks after incident peripheral neuropathy onset:<br><u>Immediate change:</u> Modify linezolid (dose reduction, temporary interruption, discontinuation) 0–48 days after peripheral neuropathy onset (Weeks 1–7)<sup>‡</sup><br><u>Deferred change:</u> Modify linezolid 49–181 days after onset (Weeks 8–26)<sup>‡</sup><br><u>No change:</u> Continue linezolid 600 mg daily within 0–181 days after onset (Weeks 1–26)<sup>‡</sup><br>We allow participants to deviate from their assigned strategy if they have any adverse event other than grade 1 or 2 peripheral neuropathy that necessitates a change to linezolid.<br>Clinicians have discretion to make other regimen changes (e.g., discontinuing cycloserine, adding new drugs) and prescribe medication for neuropathic pain. | Same. To distinguish from temporary short-term changes, we consider a strategy-defining change as one lasting $\geq 14$ consecutive days. |
| **Assignment procedure** | We randomly assign participants to one of the three treatment strategies. Participants and investigators know the assigned strategy (unblinded). | At peripheral neuropathy onset (time zero), each participant is cloned into three copies, and we assign each clone to one of the three treatment strategies. We artificially censor participant-clones at the first deviation from their assigned strategy, unless they have an allowable reason for |
|  |  | nonadherence. |
| **Follow-up** | Follow-up starts on the date of peripheral neuropathy onset (time zero) and ends at the end of MDR/RR-TB treatment (including date of death or last day in treatment if lost to follow-up). | Same. |
| **Outcome** | Successful vs. unsuccessful MDR/RR-TB treatment | Same. |
| **Causal contrasts** | Intention-to-treat effect; Per-protocol effect | Observational analog of the per-protocol effect. |
| **Identifying assumptions** | Randomization justifies no unmeasured confounding for the intention-to-treat effect.<br>For the per-protocol effect, no unmeasured confounding conditional on measured baseline covariates, and correct specification of the models used to estimate inverse probability of censoring weights. | Same as the per-protocol effect. |
| **Data analysis plan** | <u>Intention-to-treat</u>: Estimate and compare the risk (probability) of the outcome under each treatment strategy with the risk under continuing linezolid 600 mg daily, using risk differences and risk ratios. This analysis does not require adjustment for confounders.<br><u>Per-protocol</u>: Same as intention-to-treat, but we censor participants when they deviate from their assigned strategy (except for prespecified allowable reasons), and we address potential selection bias from censoring using inverse probability of censoring weighting. | Same as per-protocol, except that we use participant-clones and adjust for baseline covariates. |

\*All sites except the Democratic People's Republic of Korea due to major differences in clinical protocols related to treatment and the monitoring and reporting of adverse events.
<sup>†</sup>'Daily' refers to 6 or 7 days per week, depending on the TB program.
<sup>‡</sup>We defined Week 1 as days 0–6 from peripheral neuropathy onset, Week 2 as days 7–13, and so on, such that Weeks 1–7 correspond to days 0–48 and Weeks 8–26 to days 49–181.

Follow-up would start at the date of onset of the peripheral neuropathy adverse event (“time zero”) and end on the last day of MDR/RR-TB treatment. At time zero, we would randomly assign participants to one of three strategies determining the timing of a linezolid change (i.e., dose reduction, temporary interruption, or discontinuation) in response to peripheral neuropathy: (1) immediate change, 0–48 days after peripheral neuropathy onset (Weeks 1–7); (2) deferred change, 49–181 days after onset (Weeks 8–26); or (3) no change, 0–181 days after onset (Weeks 1–26). We defined the immediate-change window as Weeks 1–7 after peripheral neuropathy onset to capture changes to linezolid made by the next scheduled visit (typically every 4 weeks [31]). This window allows for delays of up to 3 weeks but stops short of the timeframe in which a second follow-up visit would usually occur. For immediate and deferred change strategies, clinicians would determine the specific management strategy at their discretion [32]. Dose reduction could include intermittent dosing (e.g., 600 mg three times weekly) or 300 mg daily. To ensure clinical relevance, we defined a strategy-defining change as one lasting ≥14 consecutive days. We would allow additional modifications to linezolid after adherence to the assigned strategy, and other peripheral neuropathy interventions (e.g., discontinuation of cycloserine, prescription of neuropathic pain medication) would be allowed at any time. We allowed participants to deviate from their assigned strategy if they experienced any adverse event (other than grade 1 or 2 peripheral neuropathy) that necessitated a change to linezolid, inclusive of worsening to grade ≥3 peripheral neuropathy.

We defined the primary outcome as successful MDR/RR-TB treatment, using a standardized algorithm that identified treatment failure at its earliest occurrence [35]. Successful outcomes would include cure and treatment completion; unsuccessful outcomes would include treatment failure, death, and loss to follow-up. The causal contrasts of interest would be the intention-to-treat effect and the per-protocol effect. For the intention-to-treat effect, we would estimate and compare the risk (probability) of the outcome under each treatment strategy with the risk under continuing linezolid 600 mg daily, using risk differences and risk ratios; no adjustment for confounders would be required. We would use a similar analysis plan for the per-protocol effect, except that we would censor participants when they deviated from their assigned strategy (except for prespecified allowable reasons) and address potential selection bias using inverse probability of censoring weighting, to account for the fact that censored participant-clones differed systematically from those who remained adherent to their assigned strategy.

### Target trial emulation

The emulation using data from the endTB Observational Study mainly differed from the target trial in the treatment assignment procedure. Because we could not randomize patients, we used a clone–censor–weight approach to approximate the per⍰protocol effect that the target trial would have produced. We created three copies (‘clones’) of each eligible participant’s data and assigned each clone to one of the three strategies, censoring each clone when they deviated from their assigned strategy (e.g., a clone assigned to immediate change would be censored at Week 8 if they continued linezolid beyond 48 days after peripheral neuropathy onset). We allowed participant-clones to deviate from their strategy for the same reasons as in the target trial, specifically, adverse events other than grade 1 or 2 peripheral neuropathy that necessitated a change to linezolid (including progression to grade 3–4 neuropathy). **Supplementary Figure 1** provides an overview and example of the clone–censor–weight process. Our primary estimand was the observational analog of the per-protocol effect, and data analysis followed a similar process as the per-protocol effect in the target trial with inverse probability of censoring weighting but using participant-clones. We could not meaningfully estimate an intention⍰to⍰treat effect because strategies were defined by events during follow⍰up, not by a strategy known at time zero.

## Statistical analysis

Directed acyclic graphs **(Supplementary Figure 2)** depict the analysis design, and **Supplementary Table 1** provides the rationale for including baseline and post-baseline covariates.

To estimate the denominator of the inverse probability of censoring weights, we fitted a pooled logistic regression for the probability of being uncensored at each participant⍰week (**Supplementary Table 2**), including week of follow⍰up (linear and quadratic terms), clone treatment strategy assignment, baseline covariates, and post-baseline covariates. Baseline covariates were site (Kazakhstan vs. others), number of weeks on MDR/RR⍰TB treatment (with linezolid) at peripheral neuropathy onset, earliest peripheral neuropathy adverse event grade, underweight status, presence of cavitary disease, sputum smear positivity, and whether the regimen contained bedaquiline or a fluoroquinolone. The same clinical variables (underweight, cavitary disease, sputum smear positivity, and regimen containing bedaquiline or a fluoroquinolone), together with the presence of peripheral neuropathy symptoms, were also included as post-baseline, time-varying covariates. Time⍰varying covariate values were updated or carried forward each treatment week, depending on the availability of new information. We estimated stabilized weights with a numerator using a second logistic model of being uncensored with week of follow-up (both linear and quadratic terms), clone treatment strategy assignment, and baseline covariates. **Supplementary Table 3** presents the full specification of weights and their distributions.

In the primary analysis, we used a single weighted logistic regression outcome model to estimate the probabilities of MDR/RR-TB treatment success for each treatment strategy among the uncensored participant-clones, applying stabilized inverse probability of censoring weights. We standardized the probabilities by averaging over the joint distribution of baseline covariates in the original (uncloned) cohort. We obtained 95% confidence intervals for the probabilities and their absolute differences and ratios using the bias-corrected and accelerated bootstrap with 1,000 resamples, resampling at the participant level (i.e., drawing individuals with replacement and including all of their cloned records in each replicate). We used the no-change strategy as the comparator for both immediate and deferred change strategies.

We conducted two sensitivity analyses: (1) fitting separate logistic regression models within each treatment strategy, allowing covariate effects to differ by strategy. The primary analysis offered greater precision under the assumption of no treatment–covariate interaction. If this sensitivity analysis yielded similar results to the primary analysis, the assumption of no treatment–covariate interaction would be reasonable; (2) reclassifying the outcome of loss to follow-up from an unsuccessful treatment outcome (i.e., worst-case scenario, but standard in TB treatment [6]) to a successful treatment outcome (i.e., best⍰case scenario) to assess how sensitive our conclusions are to different assumptions about the outcomes of individuals lost to follow⍰up.

We conducted all analyses using SAS version 9.4 (SAS Institute, Cary, NC) and R version 4.5.2 (R Foundation for Statistical Computing, Vienna, Austria).

## RESULTS

Of the 2,788 endTB Observational Study participants enrolled in 17 countries, 329 met inclusion criteria: they were at eligible sites, had pulmonary MDR/RR⍰TB, and initiated a regimen containing linezolid 600 mg daily. All 329 experienced peripheral neuropathy while receiving linezolid 600 mg daily in the first 6 months after cohort enrollment (**Figure 1**). We excluded 6 participants who enrolled in the cohort ≥30 days after treatment initiation and 20 participants who initially had grade ≥3 peripheral neuropathy. Our final sample included 303 participants in 12 countries: Kazakhstan (183 [60.4%]), Peru (28 [9.2%]), Armenia (24 [7.9%]), Lesotho (15 [5.0%]), Pakistan (12 [4.0%]), Georgia (12 [4.0%]), Indonesia (7 [2.3%]), Belarus (6 [2.0%]), South Africa (6 [2.0%]), Ethiopia (4 [1.3%]), Kyrgyzstan (3 [1.0%]), and Myanmar (3 [1.0%]).

**Figure 1.**
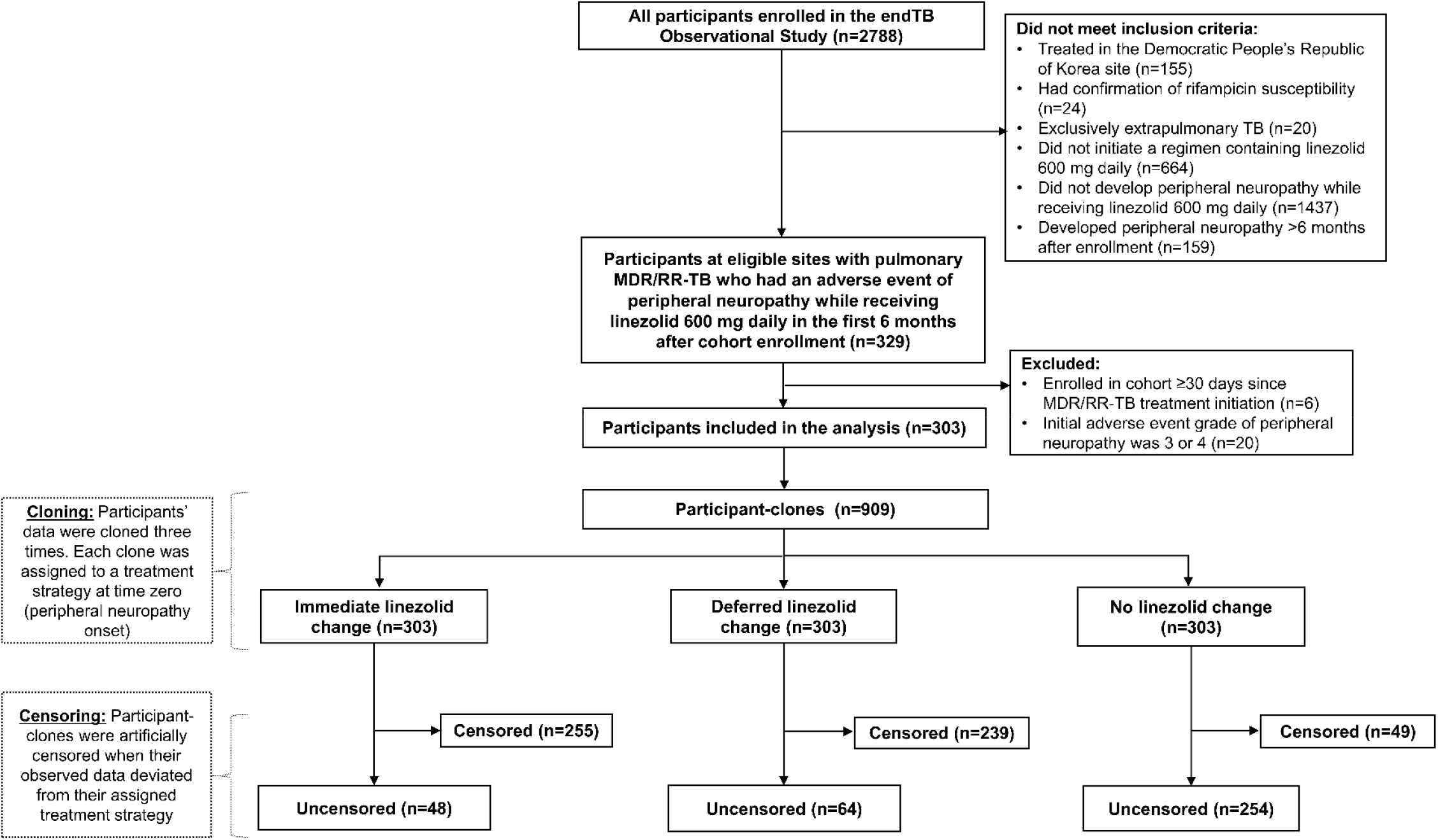
Flow diagram of inclusion into the analysis, cloning, and artificial censoring. This flow diagram shows cohort selection through inclusion and exclusion criteria, construction of the cloned data set, and the subsequent artificial censoring process. One participant did not have a TB treatment outcome evaluated, so their three participant-clones were not included in the analysis after computation of inverse probability of censoring weights. **Alt text**:Flow diagram showing cohort selection through inclusion and exclusion criteria, construction of the cloned data set, and the subsequent artificial censoring process

### Participant characteristics and incident peripheral neuropathy

Among the 303 participants, 112 (37.0%) were female, and the median (interquartile range [IQR]) age was 37 (29–47) years, with a minimum of 15 years and maximum of 74 years (**Table 2**). The analytic sample had a high burden of non-drug risk factors for peripheral neuropathy: cigarette smoking (99/299 [33.1%]), alcohol consumption (33/297 [11.1%]), HIV (27/302 [8.9%]), hepatitis C virus antibody positive (36/291 [12.4%]), diabetes (44/300 [14.7%]), and underweight (102/302 [33.8%]). At enrollment, 71/292 (24.3%) had pre-existing peripheral neuropathy.

**Table 2.** Participant characteristics at MDR/RR-TB treatment initiation (cohort enrollment)

| Characteristic | N with data | n (%) or median (IQR) |
| --- | --- | --- |
| Female sex | 303 | 112 (37.0) |
| Age in years | 303 | 37 (29–47) |
| Smokes ≥1 cigarette daily | 299 | 99 (33.1) |
| Drinks alcohol | 297 | 33 (11.1) |
| Living with HIV | 302 | 27 (8.9) |
| HCV antibody positive | 291 | 36 (12.4) |
| Diabetes | 300 | 44 (14.7) |
| Underweight (BMI <18.5 kg/m<sup>2</sup>) | 302 | 102 (33.8) |
| Pre-existing peripheral neuropathy* | 292 | 71 (24.3) |
| Bilateral lung involvement | 292 | 189 (64.7) |
| Lung fibrosis | 285 | 190 (66.7) |
| Cavitary disease | 288 | 218 (75.7) |
| Positive sputum smear | 285 | 161 (56.5) |
| Smear grade 2+ or 3+ (among positive) | 161 | 76 (47.2) |
| Positive sputum culture | 283 | 173 (61.1) |
| Prior TB treatment with second-line drugs | 278 | 248 (89.2) |
| Fluoroquinolone resistance | 286 | 190 (66.4) |
| Number of likely effective drugs in initial regimen<sup>†</sup> | 303 | 4 (4–5) |

BMI, body mass index; IQR, interquartile range; HCV, hepatitis C virus. Participant characteristics are shown for the analytic sample before cloning. In the cloned dataset, each participant is replicated under all three treatment strategies, so baseline characteristics are identical across the three strategies.
\*Symptom score >0 or past medical history of peripheral neuropathy (if the symptom score was not available).
<sup>†</sup>A drug was considered likely effective if all reported resistance testing to that drug confirmed susceptibility, or, in the absence of drug susceptibility testing, the individual had not previously received the drug for ≥1 month.

Overall, 218 (71.9%) participants had grade 1 and 85 (28.1%) had grade 2 peripheral neuropathy at onset, which occurred a median (IQR) of 11 (4–18) weeks after MDR/RR-TB treatment initiation with linezolid 600 mg daily (**Table 3**). At peripheral neuropathy onset, 92 (30.4%) participants were underweight, 221/290 (76.2%) had cavitary disease, 65/303 (21.5%) had a positive sputum smear, and 76/301 (25.2%) had a positive sputum culture. Regimens contained bedaquiline for 243/303 (80.2%) participants and a fluoroquinolone for 153/303 (50.5%) participants. In addition, 149/303 (49.2%) were also receiving cycloserine in their regimen.

**Table 3.** Participant characteristics at peripheral neuropathy onset (time zero)

| Characteristic | N with data | n (%) or median (IQR) |
| --- | --- | --- |
| Initial grade of peripheral neuropathy | 303 |  |
| Grade 1 (mild severity) |  | 218 (71.9) |
| Grade 2 (moderate severity) |  | 85 (28.1) |
| Weeks since MDR/RR-TB treatment initiation,*<br>i.e., duration on linezolid 600 mg daily | 303 | 11 (4–18) |
| Underweight<sup>†</sup> | 303 | 92 (30.4) |
| Cavitary disease<sup>†</sup> | 290 | 221 (76.2) |
| Positive sputum smear<sup>†</sup> | 303 | 65 (21.5) |
| Positive sputum culture<sup>†</sup> | 301 | 76 (25.2) |
| Regimen contained another current WHO Group<br>A drug besides linezolid |  |  |
| Bedaquiline | 303 | 243 (80.2) |
| Fluoroquinolone (levofloxacin or moxifloxacin) | 303 | 153 (50.5) |
| Regimen contained another drug that is a major<br>cause of peripheral neuropathy<sup>‡</sup> |  |  |
| Cycloserine | 303 | 149 (49.2) |
| Terizidone | 303 | 6 (2.0) |
| High-dose isoniazid | 303 | 8 (2.6) |

IQR, interquartile range; WHO, World Health Organization. Participant characteristics are shown for the analytic sample before cloning. In the cloned dataset, each participant is replicated under all three treatment strategies, so baseline characteristics are identical across the three strategies.
\*This time coincides with cohort enrollment in this analytic sample.
<sup>†</sup>Based on the latest measurement or result before the date of peripheral neuropathy onset.
<sup>‡</sup>Pyridoxine (vitamin B<sub>6</sub>) was routinely co-prescribed with these medications.

### Description of linezolid changes during follow-up

Of the 303 participants, 90 (29.7%) had a linezolid modification in the 26-week period after peripheral neuropathy onset. In Weeks 1–7 (corresponding to the immediate change strategy), 47 participants had a linezolid modification, a median (IQR) of 1 (0–4) week after peripheral neuropathy onset. Peripheral neuropathy onset was a median (IQR) of 14 (2–21) weeks after treatment initiation and the initial severity was grade 1 for 27 (57.5%) participants, with the remainder having grade 2. Of these 47 participants, 33 (70.2%) had an initial dose reduction and 14 (29.8%) had an initial interruption. The median (IQR) duration of these initial changes was 36 (8–64) weeks. Among those with a dose reduction, 20 (60.6%) changed to 300 mg daily for 6 or 7 days per week and the remainder to intermittent dosing.

In Weeks 8–26 (corresponding to the deferred change strategy), 43 participants had a linezolid modification, a median (IQR) of 13 (10–20) weeks after peripheral neuropathy onset. Peripheral neuropathy onset was a median (IQR) of 11 (3–19) weeks after treatment initiation and the initial severity was grade 1 for 30 (69.8%) participants, with the remainder having grade 2. Of these 43 participants, 23 (53.5%) had an initial dose reduction and 20 (46.5%) had an initial interruption. The median (IQR) duration of these initial changes was 27 (9–55) weeks. Among those with a dose reduction, 21 (91.3%) changed to 300 mg daily for 6 or 7 days per week and the remainder to intermittent dosing.

### End-of-treatment MDR/RR-TB and peripheral neuropathy outcomes

The median (IQR) time from peripheral neuropathy onset to the end of treatment was 75 (67–84) weeks. Excluding one person who lacked an end-of-treatment outcome, 257/302 (85.1%) participants with outcomes had a successful treatment outcome (**Supplementary Table 4**). Of the 45/302 (14.9%) with an unsuccessful outcome, 22 (7.3%) had treatment failure, 11 (3.6%) died (2 deaths were known to be TB-related), and 12 (4.0%) were lost to follow-up.

Of the 273 participants with an end-of-treatment peripheral neuropathy assessment, 172 (63.0%) were symptom free, 77 (28.2%) had mild symptoms, 20 (7.3%) had moderate symptoms, and 4 (1.5%) had severe symptoms (**Supplementary Table 5**). Participants with mild peripheral neuropathy at onset were more likely to be symptom free at the end of treatment than those with moderate neuropathy (65.7% vs. 55.6%).

### Primary analysis

An overview of censoring in the cloned dataset is shown in **Figure 1**. In the primary analysis, the weighted and standardized probabilities of successful treatment were 84.7% (95% CI: 69.2%, 92.9%) for the immediate change strategy, 78.9% (95% CI: 65.9%, 87.1%) for the deferred change strategy, and 85.2% (95% CI: 80.5%, 89.1%) for the no change strategy (**Table 4**). Compared with no change, immediate change had a –0.5 percentage-point difference in the probability of treatment success (95% CI: –14.3, 8.8) and a treatment success ratio of 0.99 (95% CI: 0.83, 1.11). Compared with no change, deferred change had a –6.3 percentage-point difference in the probability of treatment success (95% CI: –18.2, 0.8) and a treatment success ratio of 0.93 (95% CI: 0.78, 1.01).

**Table 4.** Effect of linezolid management strategy in the first 26 weeks after peripheral neuropathy onset on the probability of a successful MDR/RR-TB end-of-treatment outcome (primary analysis)

| **Linezolid management strategy** | **Weighted\* and standardized<sup>†</sup> probability of treatment success (95% CI)** | **Difference in treatment success (95% CI)** | **Treatment success ratio (95% CI)** |
| --- | --- | --- | --- |
| Immediate change | 84.7% (69.2%, 92.9%) | −0.5 (−14.3, 8.8) | 0.99 (0.83, 1.11) |
| Deferred change | 78.9% (65.9%, 87.1%) | −6.3 (−18.2, 0.8) | 0.93 (0.78, 1.01) |
| No change | 85.2% (80.5%, 89.1%) | Referent | Referent |

CI, confidence interval.
To estimate probabilities, we used a single logistic regression outcome model without interaction terms. Probabilities were rounded to the first decimal.
\*We applied stabilized inverse probability of censoring weights to uncensored participant-clones.
<sup>†</sup>We averaged predicted probabilities over the joint distribution of baseline covariates in the uncloned participant dataset: Site (Kazakhstan vs. others); number of weeks on MDR/RR-TB treatment (with linezolid) at peripheral neuropathy onset; earliest peripheral neuropathy adverse event grade; underweight status; presence of cavitary disease; sputum smear positivity; regimen contained bedaquiline; regimen contained a fluoroquinolone.

The two sensitivity analyses using separate models by treatment strategy and reclassifying loss to follow-up both yielded qualitatively similar results (**Supplementary Table 6**).

## DISCUSSION

In this target trial emulation among people receiving longer, individualized MDR/RR⍰TB regimens, immediate linezolid modification had a similar probability of treatment success as continuing linezolid 600 mg daily during the first 26 weeks after onset of mild or moderate peripheral neuropathy. Although confidence intervals were wide, these findings provide some reassurance: when clinicians needed to modify linezolid early in treatment to manage peripheral neuropathy, doing so did not appear to substantially jeopardize treatment success compared with maintaining linezolid 600 mg daily. Individuals with a deferred modification to linezolid had a modestly lower probability of treatment success compared with those who continued linezolid 600 mg daily. Any apparent difference should be interpreted cautiously. It could reflect: (1) unmeasured or residual confounding (e.g., worse prognosis among those whose linezolid was modified later); (2) differences in the type or intensity of linezolid modifications between the immediate and deferred groups; or (3) better overall tolerability in the no⍰change group that was not fully captured in our measured covariates.

To our knowledge, this is the first analysis to compare the effect of different linezolid management strategies after incident peripheral neuropathy on MDR/RR⍰TB treatment outcomes. Our findings are broadly consistent with data from other studies suggesting that linezolid 600 mg daily may not be required for the entire duration of treatment. Structured dose reductions from linezolid 600 mg daily to 300 mg daily in an open-label trial in India [36, 37] (after 9 or 13 weeks in a 26–week regimen) did not compromise efficacy while reducing the incidence of peripheral neuropathy. Additional evidence supporting structured dose reductions comes from other major trials [19, 20, 38] and an individual participant data meta-analysis [13]. In our study, most participants with an immediate change to linezolid had received a duration of linezolid 600 mg daily at peripheral neuropathy onset similar to that in these studies. However, there were major differences in regimens (i.e., longer, individualized) and patient characteristics (e.g., high burden of advanced TB disease, fluoroquinolone resistance) in our study that limit comparability.

The relevance of these findings might extend to newly endorsed all-oral shortened regimens, which all contain linezolid. Most participants in our study were receiving a regimen with at least four likely effective drugs including bedaquiline. With incident peripheral neuropathy occurring in the first six months of treatment, such a sufficiently potent regimen may have buffered against the impact of linezolid modifications. Nevertheless, generating evidence on linezolid management strategies is a high priority for these new regimens, as peripheral neuropathy remains a main treatment-limiting adverse event [16, 19-23, 39-41]. Our target trial emulation provides a methodological framework for this work. We anticipate that, because these regimens are standardized, comparative effectiveness analyses will have cleaner comparisons and require fewer assumptions regarding clinical discretion. However, future analyses on this topic require high-quality observational cohort data, underscoring the critical importance of investment in MDR/RR-TB cohort studies and routinely collected programmatic data.

The application of rigorous methods is a major strength of our study, as they improve the quality of observational analyses [28]. We also leveraged a diverse, global cohort of people treated for MDR/RR-TB with a high burden of advanced TB disease and fluoroquinolone resistance. Although the endTB Observational Study is among the largest MDR/RR-TB cohorts, our sample size for this analysis was modest. A large proportion of participants did not have a linezolid modification, which likely reflected either a watch-and-wait approach or alternative management strategies, such as modifying other drugs in the regimen or use of neuropathic pain medications, and possibly, some false-positive identification of peripheral neuropathy.

Because of the large number of participants with this treatment strategy, confidence intervals for the no-change group were relatively precise. In contrast, the smaller numbers of participants in the immediate- and deferred-change groups led to imprecise confidence intervals for those estimates. The largest site was Kazakhstan, which we considered a potential confounder and adjusted for in our analyses. Our results may have differed by site (i.e., effect modification), but the small sample size precluded meaningful stratified analyses. With our limited sample size and the use of complex individualized regimens, we were unable to compare specific clinical actions (e.g., linezolid dose reduction vs. discontinuation), which were left to clinical discretion. This is an important area for future research.

## CONCLUSION

We did not find evidence of a substantial negative impact of immediate linezolid modification on treatment success among individuals with mild or moderate peripheral neuropathy in the first six months of individualized MDR/RR⍰TB treatment. These findings support cautious linezolid adjustment, when needed, to manage non-severe peripheral neuropathy.

## Data Availability

Data may be requested through the endTB data sharing initiative (eDSI): https://endtb.org/data-sharing-initiative

## Acknowledgments

We thank Professor Miguel Hernán (Harvard T.H. Chan School of Public Health) for his valuable input in designing this analysis.

## Notes

**Potential conflicts of interest:** Matthew Romo, Allison LaHood, and Letizia Trevisi report research funding from the National Institutes of Health, with payments made to their institution. Lawrence Oyewusi, Mathieu Bastard, Helena Huerga, Uzma Khan, Catherine Hewison, Mahmud Rashitov, Nazgul Samieva, Camilo Gómez-Restrepo, Aga Krisnanda, Tinatin Kotrikadze, Andargachew Kumsa report research funding from Unitaid, with payments made to their institutions. Molly F. Franke and Kwonjune J. Seung report research funding from Unitaid and the National Institutes of Health with payments made to their institutions. Carole D. Mitnick reports research funding from Unitaid and the National Institutes of Health, with payments made to their institution, and unpaid participation on the Akagera Medicines Scientific Advisory Board. Michael L. Rich reports research funding from Unitaid and the National Institutes of Health, with payments made to their institution, and from Partners In Health and the Division of Global Health Equity, Brigham and Women’s Hospital. Dr. Rich also reports consulting fees from the World Health Organization; support for attending meetings and/or travel from the World Health Organization and Unitaid; and board membership with Pivot (Boston, MA) and Plants Earth Life (Natick, MA). Palwasha Y. Khan reports research funding from the National Institutes of Health and Unitaid, with payments made to their institution and participation on the Data Safety Monitoring Board for the TB ACT trial in Uganda. All remaining authors declare no potential conflicts of interest.

### Competing Interest Statement

Matthew Romo, Allison LaHood, and Letizia Trevisi report research funding from the National Institutes of Health, with payments made to their institution. Lawrence Oyewusi, Mathieu Bastard, Helena Huerga, Uzma Khan, Catherine Hewison, Mahmud Rashitov, Nazgul Samieva, Camilo Gomez-Restrepo, Aga Krisnanda, Tinatin Kotrikadze, Andargachew Kumsa report research funding from Unitaid, with payments made to their institutions. Molly F. Franke and Kwonjune J. Seung report research funding from Unitaid and the National Institutes of Health with payments made to their institutions. Carole D. Mitnick reports research funding from Unitaid and the National Institutes of Health, with payments made to their institution, and unpaid participation on the Akagera Medicines Scientific Advisory Board. Michael L. Rich reports research funding from Unitaid and the National Institutes of Health, with payments made to their institution, and from Partners In Health and the Division of Global Health Equity, Brigham and Womens Hospital. Dr. Rich also reports consulting fees from the World Health Organization; support for attending meetings and/or travel from the World Health Organization and Unitaid; and board membership with Pivot (Boston, MA) and Plants Earth Life (Natick, MA). Palwasha Y. Khan reports research funding from the National Institutes of Health and Unitaid, with payments made to their institution and participation on the Data Safety Monitoring Board for the TB ACT trial in Uganda. All remaining authors declare no potential conflicts of interest.

### Funding Statement

The funder of the endTB project is Unitaid. Bedaquiline was donated by Janssen to the Global Drug Facility and delamanid was donated by Otsuka for initial participants enrolled in the endTB Observational Study. These companies did not have any role in study design, data analysis, data interpretation, or manuscript writing. This secondary analysis was funded entirely by the National Institute of Allergy and Infectious Diseases of the US National Institutes of Health (R01AI146095, funding to Molly F. Franke, Carole D. Mitnick, Michael L. Rich, Kwonjune J. Seung, Letizia Trevisi, Matthew Romo). The content is solely the responsibility of the authors and does not necessarily represent the official views of the National Institutes of Health.

### Author Declarations

This study involves human participants and the endTB Observational Study protocol was approved by ethics committees for each consortium partner (Partners Healthcare: #2015P001669; IRD: #IRD_IRB_2015_08_001; MSF: #1550) and in each country. Participants (or a guardian if the individual was a minor, as defined by local legal requirements) provided written informed consent and older minors provided assent.

### Summary of Updates

We revised the manuscript based on peer reviewer comments.

