## Supplementary Files for "Comparison of three linezolid management strategies for peripheral neuropathy in multidrug- or rifampicin-resistant tuberculosis treatment: a target trial emulation"

**Supplementary Figure 1. Steps to the clone-censor-weight approach with an illustrative example**

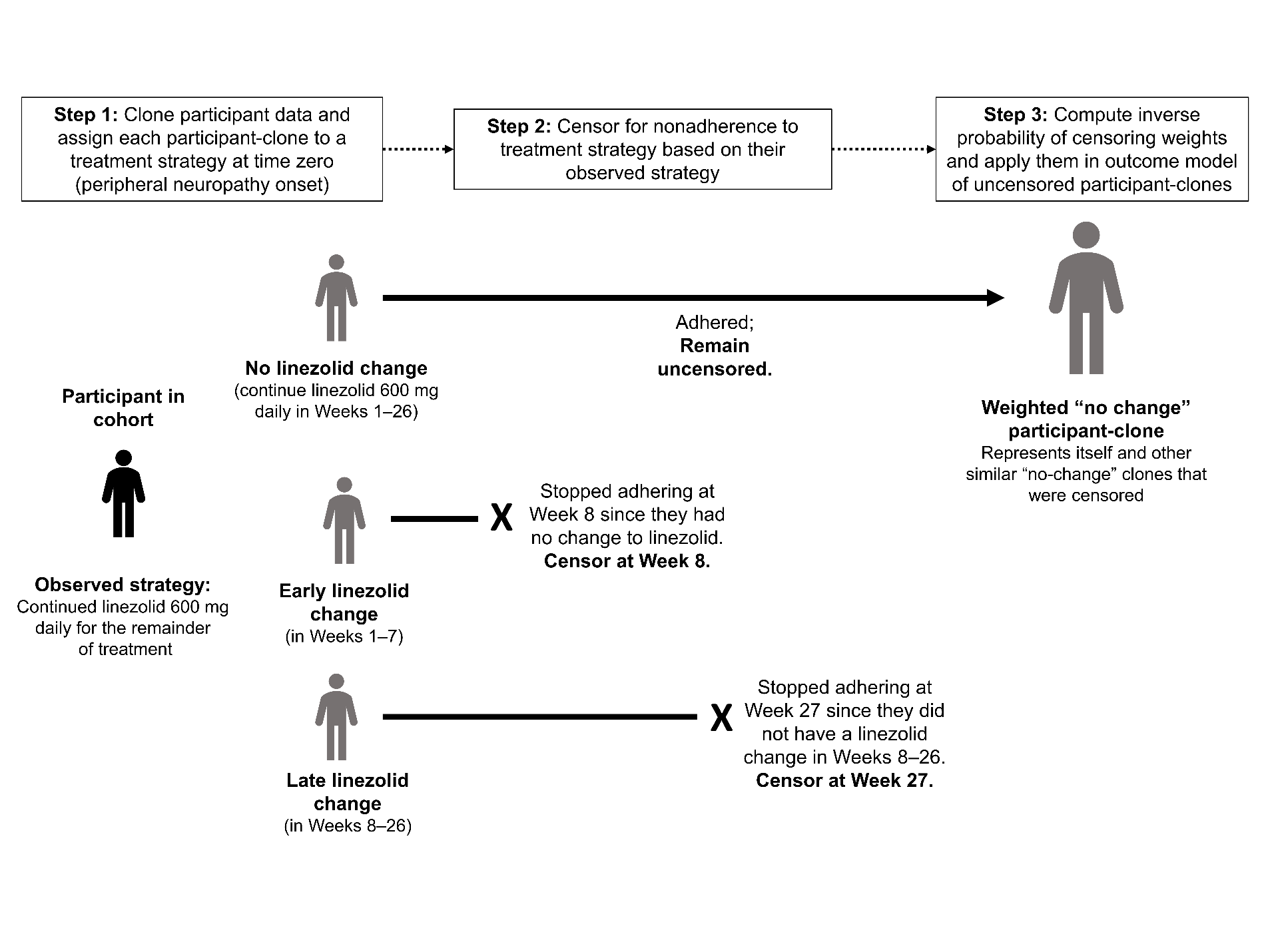

We cloned (copied) each participant’s data three times and assigned each participant‑clone to one of the three treatment strategies (**Step 1**). If a clone became non-adherent to its assigned strategy (i.e., the clone’s observed data was no longer compatible with the assigned strategy), we “artificially” censored that participant‑clone (**Step 2**), unless the nonadherence resulted from an adverse event other than grade 1 or 2 peripheral neuropathy. We computed inverse probability of censoring weights using baseline and time‑varying variables related to both censoring and the treatment outcome, and applied these weights in an outcome model of the uncensored participant‑clones (**Step 3**).

**Supplementary Figure 2. Directed acyclic graphs showing the relationship between participant-clone treatment strategy assignment and MDR/RR-TB treatment success**

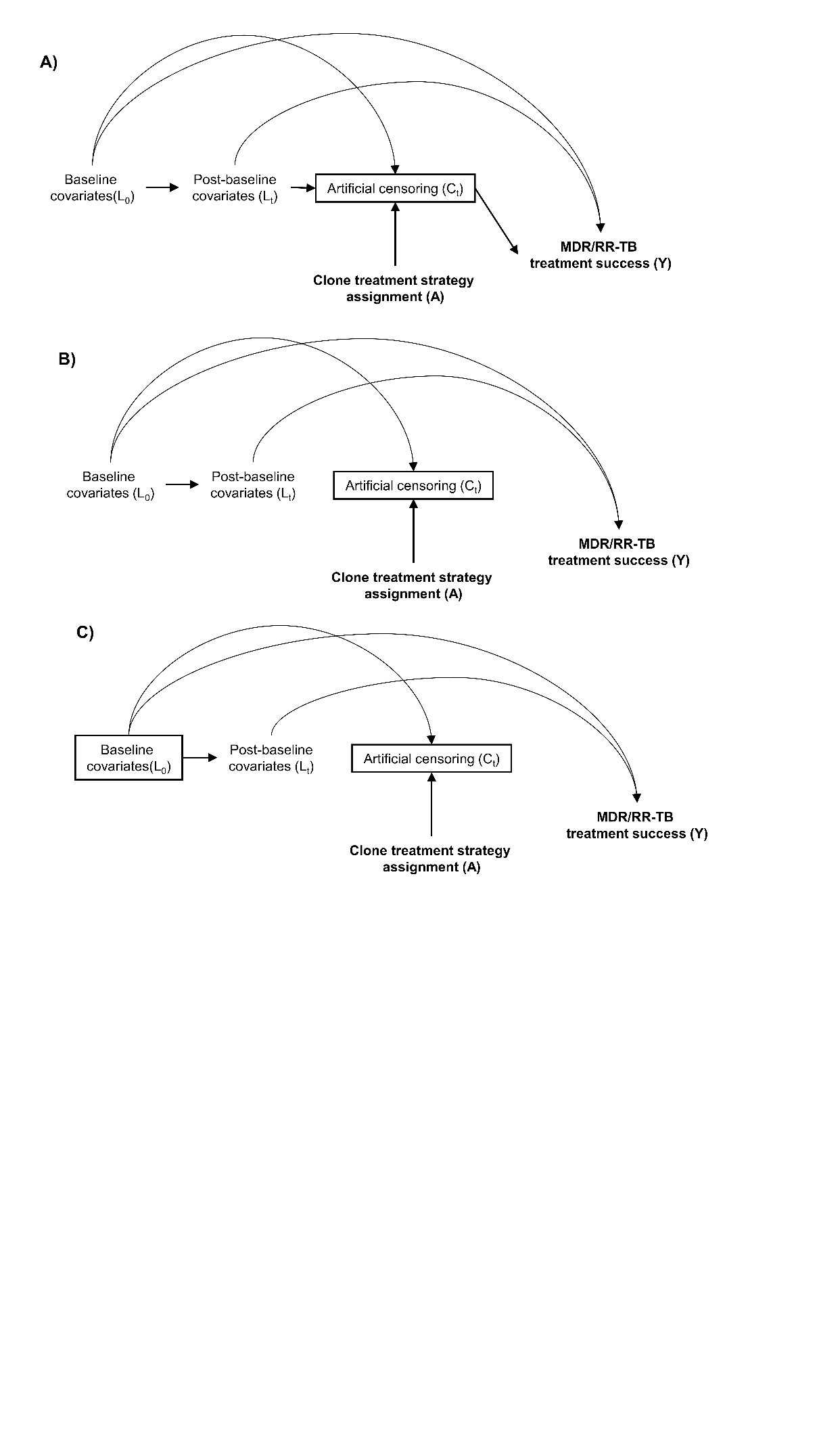

**Panel A** depicts an analysis estimating the effect of A on Y, conditioning on C_t_ (i.e., limiting the analyses to uncensored participant clones). This opens up pathways from A to Y via C_t_, L_t_, and L_0_.

**Panel B** depicts an analysis estimating the effect of A on Y, conditioning on C_t_ (i.e., limiting the analyses to uncensored participant clones), but also applying stabilized inverse probability of censoring weights. This blocks pathways from A to Y via C_t_ and L_t_. Because the inverse probability of censoring weights are stabilized on L_0_, the arrow from L_0_ to C_t_ remains.

**Panel C** depicts an analysis estimating the effect of A on Y, conditioning on C_t_ (i.e., limiting the analyses to uncensored participant clones), applying stabilized inverse probability of censoring weights, and additionally adjusting for L_0_. This blocks the final open pathway from A to Y via L_0_.

**Supplementary Table 1. Rationale for covariate selection**

| **Covariate** | **When measured*** | **Rationale** |
| --- | --- | --- |
| Number of weeks on MDR/RR-TB treatment (with linezolid) | Baseline | Longer duration of linezolid received (vs. less):   - More likely to modify linezolid immediately - Increased probability of treatment success |
| Initial severity of peripheral neuropathy (grade 1 or 2) | Baseline | Grade 2 (moderate severity) peripheral neuropathy (vs. grade 1 [mild severity]):   - More likely to modify linezolid immediately - Decreased probability of treatment success (e.g., through pathways like loss to follow-up) |
| Presence of peripheral neuropathy symptoms | Post-baseline | Persistent symptoms (vs. resolution of symptoms)   - More likely to modify linezolid - Decreased probability of treatment success |
| Cavitary disease | Baseline + post-baseline | Cavitary disease (i.e., more severe TB disease; vs. no cavities):   - Less likely to modify linezolid - Decreased probability of treatment success |
| Sputum smear positivity | Baseline + post-baseline | Positive sputum smear (i.e., TB not yet responding to treatment; vs. negative)   - Less likely to modify linezolid - Decreased probability of treatment success |
| Underweight status | Baseline + post-baseline | Underweight (vs. normal weight or higher)   - Less likely to modify linezolid (due to worse prognosis), or possibly more likely (due to the perception that less linezolid is required) - Decreased probability of treatment success |
| Regimen contains bedaquiline | Baseline + post-baseline | Stronger regimen (e.g., contains bedaquiline)   - More likely to modify linezolid (i.e., increased flexibility to do so, if needed) - Increased probability of treatment success |
| Regimen contains a fluoroquinolone | Baseline + post-baseline | Same as above; additionally, people receiving a fluoroquinolone also likely have fluoroquinolone-susceptible MDR/RR-TB, strengthening the rationale |
| Site (Kazakhstan vs. all other countries) | Baseline | Predominance of data from Kazakhstan and its role as a high‑volume, clinically experienced site:   - More likely to modify linezolid - Increased probability of treatment success |

*Covariates are prognostic factors expected to be associated with linezolid modification at the onset of peripheral neuropathy (baseline) or over time (post-baseline) and MDR/RR-TB treatment success. Post-baseline covariates were treated as time varying, whereby values were updated or carried forward, depending on the availability of new information, each treatment week.

**Supplementary Table 2. Probabilities of being uncensored for each treatment strategy and follow-up period**

| **Treatment strategy assigned to participant-clone** | **Week of follow-up (week defined as the end of every 7-day period from time zero)** | | | | |
| --- | --- | --- | --- | --- | --- |
|  | **Weeks 1–7 (days 0–48)** | **Week 8 (days 49–55)** | **Weeks 9–26 (days 56–181)** | **Week 27 (days 182–188)** | **Week 28–end of treatment (days 189 onward)** |
| Immediate linezolid change | Equal to 1 | 1 − Probability of remaining on linezolid 600 mg daily through the end of Week 7 | Equal to 1 | Equal to 1 | Equal to 1 |
| Deferred linezolid change | Probability of remaining on linezolid 600 mg daily or changing linezolid only due to another adverse event* during Weeks 1–7 | Equal to 1 | Equal to 1 | 1 − Probability of remaining on linezolid 600 mg daily through Week 26 (without a linezolid change between Weeks 8 and 26) | Equal to 1 |
| No linezolid change | Probability of maintaining linezolid 600 mg daily or changing linezolid only due to another adverse event* during Weeks 1–7 | Probability of maintaining linezolid 600 mg daily or changing linezolid only due to another adverse event* during Week 8 | Probability of maintaining linezolid 600 mg daily or changing linezolid only due to another adverse event* during Weeks 9–26 | Equal to 1 | Equal to 1 |

We calculated the probability of remaining uncensored for each week starting at Week 1 (the week including time zero, i.e., peripheral neuropathy onset). For any given week, we obtained the probability that a participant‑clone remained uncensored (i.e., adhered to the assigned strategy) by taking the cumulative product of the conditional probabilities of remaining uncensored in each preceding week, including the current week. We then computed stabilized inverse probability of censoring weights (**Supplementary Table 3**) and retained the final weight for participant‑clones that remained uncensored.

*Any adverse event (other than grade 1 or 2 peripheral neuropathy) documented as the reason to change linezolid, including worsening to grade 3 or 4 peripheral neuropathy. If other adverse events were documented with grade 1 or 2 peripheral neuropathy (e.g., myelosuppression), we considered these non-neuropathy adverse events as the primary reason for modifying linezolid and would not trigger censoring.

**Supplementary Table 3. Specification and distribution of stabilized inverse probability of censoring weights**

| **Specification** | | | | | | | |
| --- | --- | --- | --- | --- | --- | --- | --- |
| **Numerator** | Treatment week (since time zero); quadratic term for treatment week; clone treatment strategy assignment; baseline covariates: site (Kazakhstan vs. others); number of weeks on MDR/RR-TB treatment (with linezolid) at peripheral neuropathy onset; earliest peripheral neuropathy adverse event grade; underweight status; cavitary disease; sputum smear positivity; regimen contained bedaquiline; regimen contained a fluoroquinolone. | | | | | | |
| **Denominator** | Treatment week (since time zero); quadratic term for treatment week; clone treatment strategy assignment; baseline covariates (as listed above for numerator); and post-baseline covariates (cavitary disease; underweight status; sputum smear positivity; regimen contained bedaquiline; regimen contained a fluoroquinolone; peripheral neuropathy symptoms present). | | | | | | |
| **Distribution** | | | | | | | |
|  | **N** | **Sum** | **Mean** | **SD** | **Median** | **IQR** | **Min, Max** |
| Immediate linezolid change | 48 | 49.5 | 1.03 | 0.39 | 0.97 | 0.75, 1.26 | 0.37, 2.16 |
| Deferred linezolid change | 64 | 62.4 | 0.97 | 0.13 | 0.98 | 0.88, 1.05 | 0.67, 1.26 |
| No linezolid change | 254 | 254.2 | 1.00 | 0.03 | 1.00 | 0.98, 1.02 | 0.93, 1.08 |
| **Overall** | 366 | 366 | 1.00 | 0.15 | 1.00 | 0.97, 1.02 | 0.37, 2.16 |

IQR, interquartile range; SD, standard deviation.

We carried the last observation forward for time-varying variables. Thirteen (4.3%) participants were missing some chest radiography data; therefore, in addition to variables for presence of cavitary disease (yes/no), we included an indicator variable for chest radiography data being available.

**Supplementary Table 4. MDR/RR-TB end-of-treatment outcome for all participants**

| **Outcome** | **All participants (N=302)*** |
| --- | --- |
| **Successful, n (%)** | **257 (85.1)** |
| Cured, n (%) | 248 (82.1) |
| Completed, n (%) | 9 (3.0) |
| **Unsuccessful, n (%)** | **45 (14.9)** |
| Died, n (%) | 11 (3.6) |
| TB-related, n | 2 |
| Not TB related, n | 7 |
| Unknown, n | 2 |
| Treatment failure, n (%) | 22 (7.3) |
| Lost to follow-up, n (%) | 12 (4.0) |

*One participant did not have an end-of-treatment outcome evaluated and was not included in the denominator.

**Supplementary Table 5. End-of-treatment peripheral neuropathy severity for all participants**

| **End-of-treatment peripheral neuropathy severity*** | **Overall (N=303)** | **Initial severity grade 1 (N=218)** | **Initial severity grade 2 (N=85)** |
| --- | --- | --- | --- |
| Grade 0 (no symptoms), n (%) | 172 (63.0) | 132 (65.7) | 40 (55.6) |
| Grade 1 (mild), n (%) | 77 (28.2) | 58 (28.9) | 19 (26.4) |
| Grade 2 (moderate), n (%) | 20 (7.3) | 9 (4.5) | 11 (15.3) |
| Grade 3 (severe), n (%) | 4 (1.5) | 2 (1.0) | 2 (2.8) |
| No assessment, n | 30 | 17 | 13 |

*Follow-up assessment closest to the date of the end of treatment, ±3 months.

**Supplementary Table 6. Sensitivity analyses for the effect of linezolid management strategy on a successful MDR/RR-TB end-of-treatment outcome**

| **Sensitivity analysis** | **Linezolid management strategy** | **Weighted and standardized probability of treatment success (95% CI)** | **Treatment success ratio (95% CI)** |
| --- | --- | --- | --- |
| Separate logistic regression models within each treatment strategy | Immediate change | 83.0% (68.6%, 96.5%) | 0.97 (0.80, 1.13) |
|  | Deferred change | 78.4% (65.5%, 90.3%) | 0.92 (0.78, 1.06) |
|  | No change | 85.3% (80.6%, 89.3%) | Referent |
| Participants lost to follow-up are reclassified as having successful treatment outcome (i.e., best-case scenario) | Immediate change | 90.5% (76.4%, 97.1%) | 1.02 (0.86, 1.10) |
|  | Deferred change | 83.3% (71.1%, 90.8%) | 0.93 (0.81, 1.01) |
|  | No change | 89.1% (85.3%, 92.7%) | Referent |

CI, confidence interval. Probabilities were rounded to the first decimal.
